# Planning optimal sequence for arteriovenous fistula creation in hemodialysis patients, using quality-of-life adjusted access patency

**DOI:** 10.1101/2023.04.06.23288219

**Authors:** Manoj Kumar, Deepa Usulumarty, Ravi Brahmbhatt, Narayan Rangaraj, Santosh Noronha, Viswanath Billa

## Abstract

**Objective:** The study proposes a decision strategy for the choice of arm for placement of an arteriovenous fistula (AVF) in patients with kidney failure who undergo hemodialysis (HD). The possible surgical sites for placement of the AVF are located in the arm and at two locations in each arm, the wrist (distal AVF) and the elbow (proximal AVF). Recommendations are made for the location of the first AVF and subsequent ones, as needed.

**Method:** A retrospective analysis of AVFs created between 2015 and 2022 in patients at five HD centers was performed. The study uses quality-adjusted survival, parametric, and nonparametric survival methods. We conducted two surveys to assess the HD patient’s quality of life. In addition, the Cox proportional hazard model is used to suggest a patient-specific strategy. The survival functions are compared using the log-rank and Tarone-Ware tests.

**Results:** The results of the multivariate analysis showed that placing AVFs on the non-dominant arm leads to superior patient quality of life compared to the dominant arm. The quality-adjusted survival was also found to be better when AVFs were located on the non-dominant arm. Considering both these aspects and also clinical constraints on the sequence, the optimal sequence is found to be non-dominant distal followed by non-dominant proximal locations, followed by a similar sequence on the dominant hand if required.

**Conclusion:** The study identifies criteria for data-driven decision-making as to good locations for AVFs for Hemodialysis and supports clinical experience and practice in this matter.

## 1. Introduction

Patients with end-stage renal disease (ESRD) need dialysis on a regular basis till transplant options become available. Hemodialysis (HD) is a commonly used procedure that requires stable vascular access. These could be of several types, such as; a central venous catheter (temporary and permanent catheter), arteriovenous fistula (AVF), and arteriovenous graft (AVG) [1]. An arterio-venous fistula is a surgical procedure to connect an artery with a vein in the arm of a patient requiring HD. Several studies have highlighted the use of arteriovenous fistulas for hemodialysis because they have fewer complications and longer patency compared with other vascular access options [1, 2].

It is observed that AVF sometimes matures sub-optimally and therefore is unusable for dialysis. AVFs should mature fully to be able to be used for prolonged periods. The average time for AVF maturation is four to six weeks. Factors associated with AVF maturation have been reviewed in the literature [3]. The optimal time to refer Chronic Kidney Disease (CKD) patients for AVF placement is crucial and should be several months before the need to start dialysis, as it takes several weeks to mature [4, 5]. A Markov model with decision trees was presented to determine the cost-effectiveness of AV fistula versus AVG [6].

The AVF can be created on any hand, at the wrist (radio-cephalic or distal), or at the elbow, known as brachio-cephalic (proximal). A proximal AVF is possible if the distal option has already been used for an earlier AVF, but the reverse is not possible for clinical reasons. The blood flow in the downstream vein at the wrist reduces after constructing a proximal AVF. Therefore, nephrologists try to construct the first AVF at the wrist to keep the upstream option open for subsequent AVFs if needed, as shown in Figure 1. The patency of different sites, such as distal and proximal, has been studied in the literature but is not specific to the arm used for fistula placement [7, 8]. The suitability of the arm is assessed before performing the AVF surgery as per standard protocol. Most of the time, the patient’s non-dominant hand is preferred over the dominant hand with the belief that this approach would avoid injury to the AVF that a dominant arm can be more prone to. However, the validation of this choice of the non-dominant arm has not been established.

**Figure 1:**
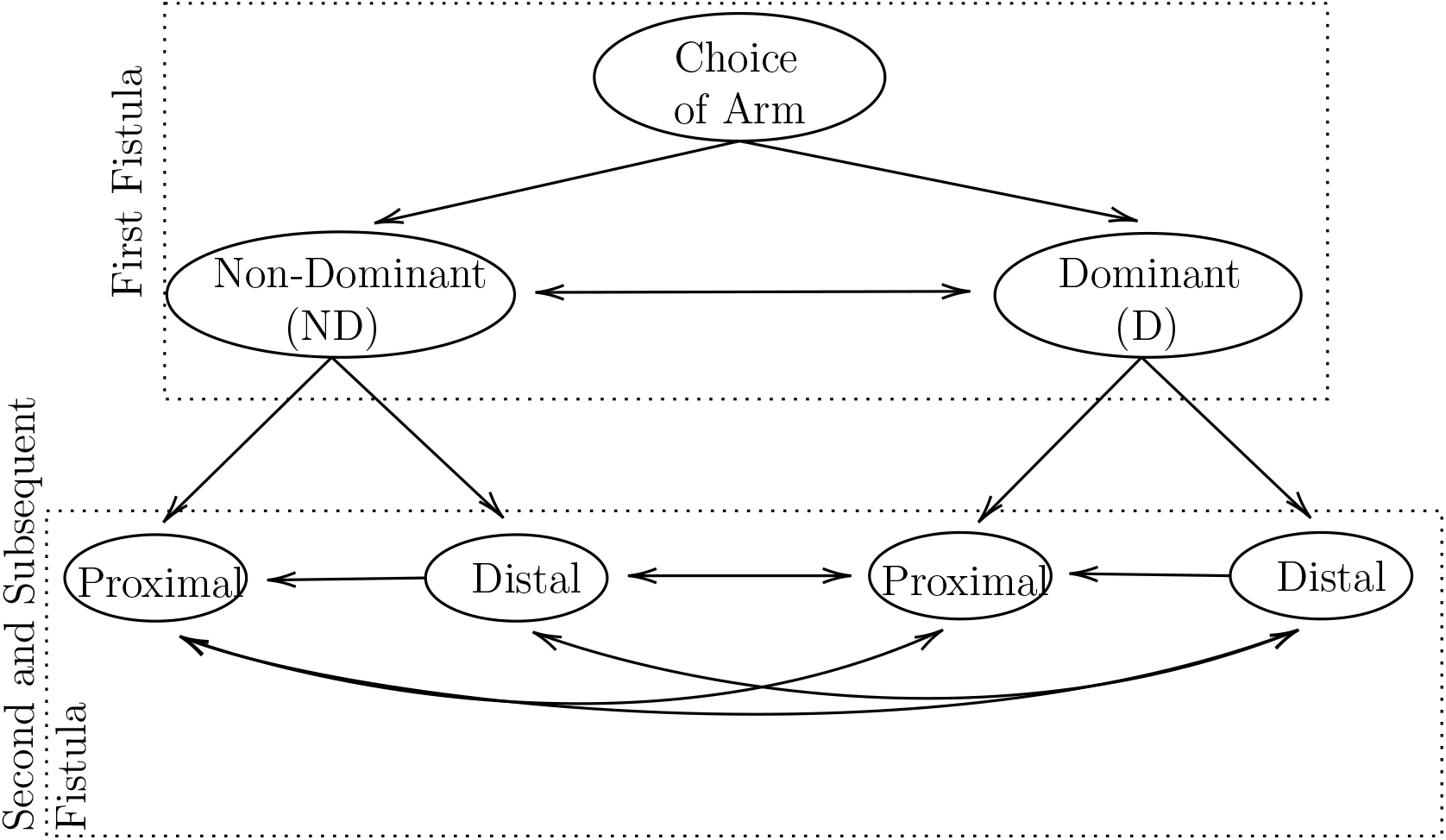
Decision flow for arteriovenous fistula placement. The arrows show the possible flow of decisions for AVF placement.

The AVF, as a vascular access option, has good survival rates. Failure modes are (i) immediate failure after construction, which is rare, (ii) suboptimal maturation, called late primary failure; and (iii) secondary failure after normal maturation. Analysis of survival statistics is in sections 4 and 6. One study highlighted the reasons and presented the guidelines for how to intervene to avoid AVF failures [2]. A decision support model for predicting graft survival in renal transplant recipients was presented [9]. Recently, a patient-specific computational model has been developed and validated in a clinical trial to predict pre-operatively the blood flow volume (BFV) in AVF for different surgical configurations on the basis of demographic, clinical, and doppler ultrasound data [10]. Although the authors have presented a model to measure the BFV pre-operatively for different configurations, they have not discussed the sequence of sites and the consequence of AVF placement at a particular site.

Quality-adjusted survival is widely used to compare different disease treatments [11, 12, 13, 14]. A widely used method is Quality-adjusted time without disease symptoms and toxicity (Q-TWiST), where TWisT health state will have a higher utility than other health states.

Various studies have been done to understand AVF patency, but none talks about the difference in patency rates between the two arms and the optimal sequence of creation of multiple AVFs. In this study, we attempt a decision plan for the creation of multiple AVFs in a sequential manner. The policy depends on the patency estimates for any AVF and the patient’s quality of life. In addition to the optimal policy for the first AVF and the optimal sequence for multiple AVFs, a contribution to this is a rigorous definition of a patient’s quality of life impacted by the AVF creation. Comparing a patient’s quality of life becomes very important in decision-making when the patency of two treatments is almost the same.

## 2. Problem Description

Hemodialysis patients require stable vascular access for prolonged dialysis. The various possible sites and their consequences have already been discussed in the introduction section 1.

For an AVF placement, there are four potential locations, viz. Dominant arm Distal, Dominant arm Proximal, Non-dominant arm Distal, and NonDominant arm Proximal.

Part one of this study proposes a threshold policy to choose the dominant or non-dominant arm for the first AVF, keeping in mind the patient’s quality of life and survival of the fistula.

The second part of the study extends the analysis to the sequences adopted when multiple AVFs were created, as hemodialysis patients usually need more than one AVF during their lifetime on dialysis. The possible clinical sequences are as follows and also depicted by the arrows in Figure 1:

1. D Distal *→* D Proximal *→* ND Distal *→* ND Proximal

2. D Distal *→* ND Distal *→* D Proximal *→* ND Proximal

3. ND Distal *→* ND Proximal *→* D Distal *→* D Proximal

4. ND Distal *→* D Distal *→* ND Proximal *→* D Proximal

A Distal AVF creation is not possible if a proximal AVF has been constructed in the first instance on either of the arms. Currently, the clinical choice and practice are to choose the ND arm distal location as the first choice for AVF. However, it needs data-driven validation, which is in this paper. The risk of (i) an accidental injury and (ii) inconvenience are both perceived to be higher for D Distal over ND distal. The patency for any AVF is improved by physical activity, which in turn enhances the blood circulation to that AVF. As the dominant hand is involved in everyday physical activities, patients do not require to undertake an additional physical effort to improve this blood circulation, as compared to the non-dominant hand.

The choice of placement of the second fistula is less clearly defined in current practice and is an open question. Therefore, we compare the four sequences implied by Fig. 1 for cases where multiple AVFs are used.

Also, after AVF surgery, patients may have challenges in performing their daily activities with that arm. Therefore, the AVF should be constructed where it would have a higher chance of survival than other potential sites and minimally impact the patient’s physical ability to perform daily activities. One of the research problems we tackle in this article is how to measure the effect of AVFs on a patient’s everyday physical activities. We define the quality of life of a patient with AVF on either arm as the weighted quantity of the effects of AVF on various activities. Therefore, it is of interest to hypothesize that the average quality of life of a patient is different in the different arms.

## 3. Methodology

This study has included various methodologies in computing quality-adjusted survival for either arm. We divide our computations into three parts; quality of life when fistula was constructed on D and ND arm, patency estimate of D and ND arm fistula, and quality-adjusted survivals. Each of these parts is elaborated on in the following subsections.

### 3.1. Computation of quality of life

AVF affects a patient’s quality of life because he or she is limited in some daily activities. We assume that the extent to which this occurs (as a factor) remains constant throughout the life of the fistula. The quality of life of a patient undergoing hemodialysis depends on many factors, such as the patient’s and AVF’s health. Here, we attempt to quantify the factor by which the quality of life of a patient is affected based on the site and location of the AVF (whether it is on the D or ND hand). It is also important to record how much difficulty a patient has in performing each activity. Moreover, each activity has a different importance in our daily life. Therefore, we propose that quality of life is a composite quantity consisting of physical, symptomatic, and psychosocial factors. Each of these factors consists of 5-6 sub-factors, e.g., physical activities include holding/gripping, stretching, rotation, pushing, constriction/pressure, and complex activities, as shown in Table 3. The quality of life is defined as the weighted sum of the impact of AVF on each activity. The weights reflect the relative importance of each activity in our everyday life.

We define the quality of life, *Q*, as:

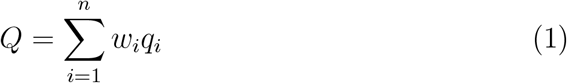

*Q* is measured on a scale from 0 to 1, where 1 indicates the highest possible quality for the patient. *w*_*i*_ and *q*_*i*_ are the relative importance and quality gain of activity *i, n* is total activities considered in the study.

We hypothesized the following to shed light on the effects of AVF on each activity:

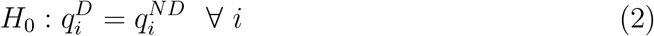

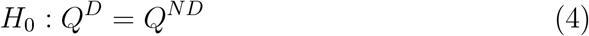

where, 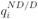 denotes the impact of ND/D AVF on *i*^*th*^ activity.

For the quality of life with respect to ND/D hand, we designed our hypothesis as follows:

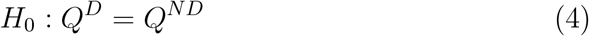

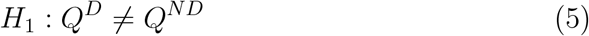

### 3.2. Survival Function

The survival function is that a subject of interest will survive past a certain time. In reliability studies, survival is a widely used term to estimate lifespan, whereas, in nephrology, the term patency is often used [15, 16].

Therefore, AVF patency is synonymous with survival in this article. The survival function can be estimated using non-parametric and parametric methods. The Kaplan-Meier survival estimate is a non-parametric method that estimates the survival probabilities using the observed survival times [17]. Non-parametric methods have advantages over the parametric method, but it is difficult to extrapolate the survival analysis beyond the study period.

Parametric methods make assumptions about the data distribution, and frequently used standard distributions are exponential, Weibull, log-normal, and log-logistic. The best parametric distribution for survival estimation is obtained based on the goodness of fit, which can be accessed using the Akaike Information Criterion (AIC). In this study, we have used all these four parametric distributions and selected the best survival function with the lowest AIC score.

The different statistics, such as the log-rank test, Wilcoxon, Tarone-Ware, and Peto-Peto, were used to compare two survival functions [18]. The median survival statistics have been presented here for all the survival functions computed in this study.

### 3.3. Quality-adjusted Patency

Quality-adjusted survival (*S*_*Q*_) of a subject is defined as the measure of the effectiveness of an intervention with a trade-off between quality of life and patency [13, 14]. Quality-adjusted survival in this article refers to quality-adjusted AVF patency rates.

In our study, we have one state that is a patient under dialysis with AVF vascular access where AVF could be on either hand. Therefore, we define it as follows: Let *S*^*i*^(*t*) denote the survival function of an AVF constructed on *i* = *{D, ND}*, represents dominant (D) and non-dominant (ND) hand, *T* denote the time horizon for which we are planning the fistula placement, and *Q*^*i*^ denotes utility of fistula on the *i* arm.

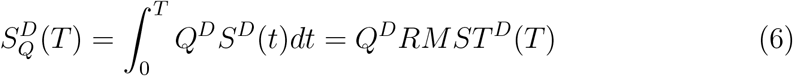

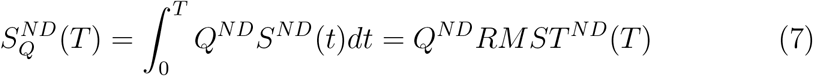

where *RMST* (*T*) refers to restricted mean survival till time *T*. It estimates the time spent in that health state.

#### Policy 1.

*The decision policy for the planning horizon, T is:*

1. *Dominant arm should be preferred if* 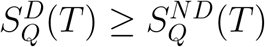.
2. *Non-Dominant arm should be preferred if* 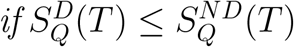.

## 4. Data Description

For this study, the AVF survival data^1^ were collected from five dialysis centers. The patient’s details are the arm used for the fistula and the time of failure of the fistula. An AV fistula can have two types of failures: primary and secondary. Primary failure is defined as the failure of the fistula within three months of its construction. The secondary failure is defined as the failure of AVF that survives beyond three months.

Figure 2 shows the details of the data used in this analysis. Out of 43 primary AVF failures, 12 (28%) AVFs were on the D hand, and the rest were on the ND hand. The radio-cephalic (distal) fistulas had higher primary failure than brachiocephalic (proximal) fistulas for both hands.

**Figure 2:**
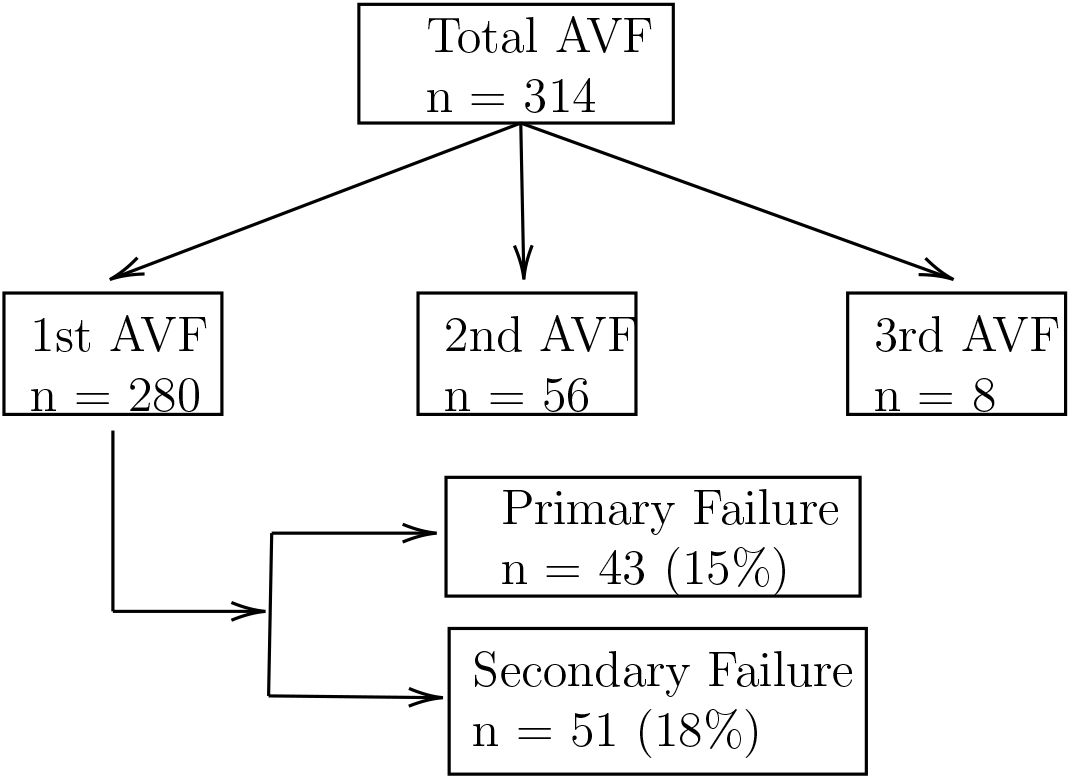
Data Description.

**Figure 3:**
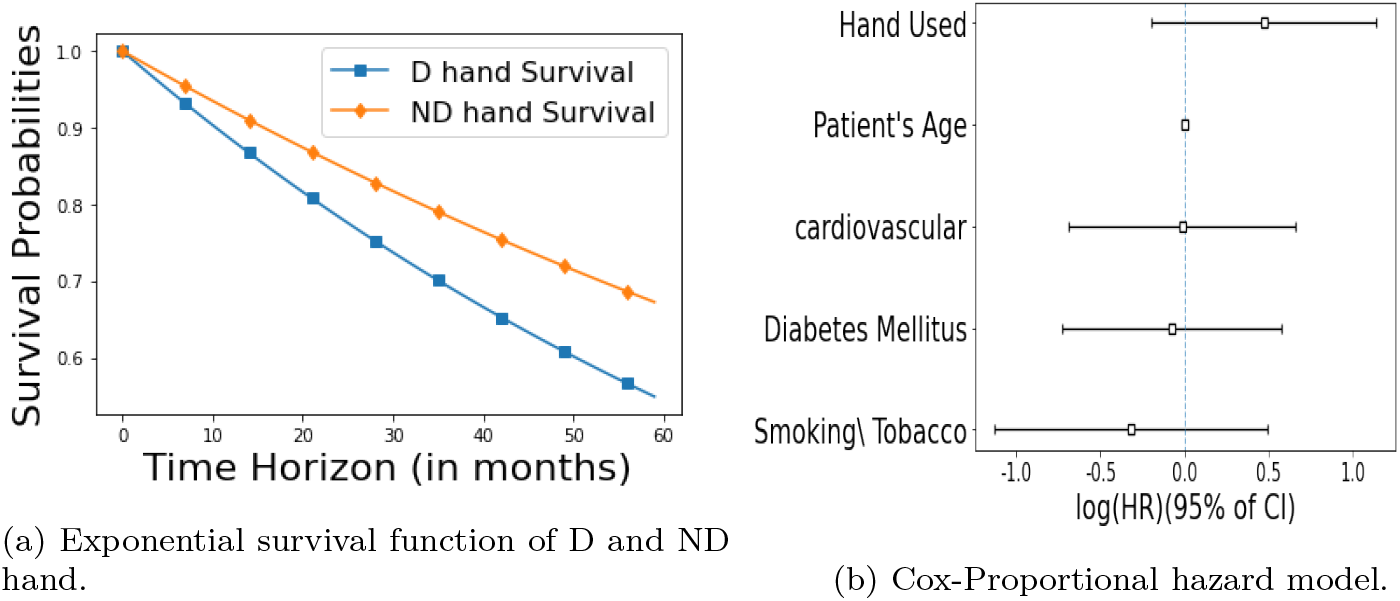
Univariate and Multivariate survival of dominant and non-dominant hand.

**Figure 4:**
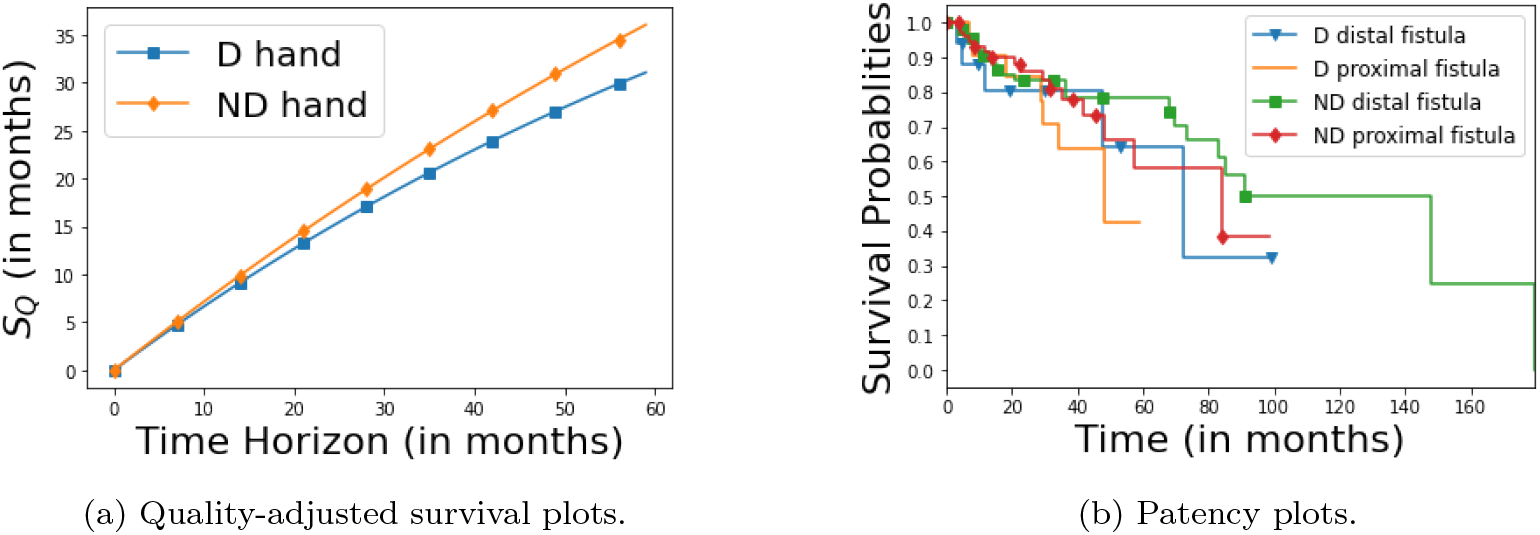
Quality adjusted survivals for D and ND hand for first AVF and Kaplan Meier estimates of different AVF based on locations.

We conducted two different surveys for quality-of-life computation. One included questions related to the relative importance of various activities in our daily life, and the other one is related to the impact of AVF on everyday activities. We designed our questionnaire to quantify the impact of AVF based on the previous research [19]. The questionnaire listed various physical activities of daily living like rotation, pushing, and complex activities with more than one type of motion and the degree of difficulty in performing them. It also included clinical symptoms like pain, weakness, stiffness, and anxiety.

## 5. Quality Computation Results

The survey for the impact of AVF on daily activities was conducted and responses were received from 127 patients (25 had AVF on dominant and 102 had it on non-dominant). The multivariate analysis of variance (MANOVA) test was performed to see the difference between the dominant and non-dominant arm groups. We found a statistically significant difference (*p ≈* 0.00) between the two groups, implying that our null hypothesis (2) had to be rejected. Although the quality gain of each activity is higher for the non-dominant arm group as compared to the dominant arm group, the Bonferroni correction method showed that none of the activities were statistically significant in a univariate analysis at a 5% significance level as shown in Table 3. Though we do not have statistical evidence of AVF’s consequences for each activity, it has statistical impact on the patient’s daily life for hands.

This type of survey can not be administered to each patient who needs dialysis. It is a one-time collection of the impact of different factors and the consequence of constructing a fistula on either hand. The aggregate response is used as a general guideline for fistula constructions. Any patient-specific information would then be used over and above this.

The importance of one activity to a person may be higher (equal or lower) than that of another. The relative importance of specific activities for a given person may vary, which confuses a nephrologist’s decision-making process. Our approach consolidates individual preferences in the order of activities to derive relative importance.

All 18 activities were divided into physical, symptomatic, and psychosocial factors. We conducted a survey for the relative importance of each of the activities under physical factors. The respondents were requested to rate these activities per their personal preference (scale of 1 to 9). We interviewed the nephrologist for the activities under symptomatic and psychosocial factors and asked them to rate these on a scale of one to nine. On a rating scale, one is “ not important”, and nine is “ very important.” We have also asked nephrologists to rate these three categories among themselves on a scale of one to three, where an ascending order of these numbers refers to increasing importance. We randomly generated 1000 samples for each category using a uniform distribution with the means taken from Table 2.

**Table 1:**
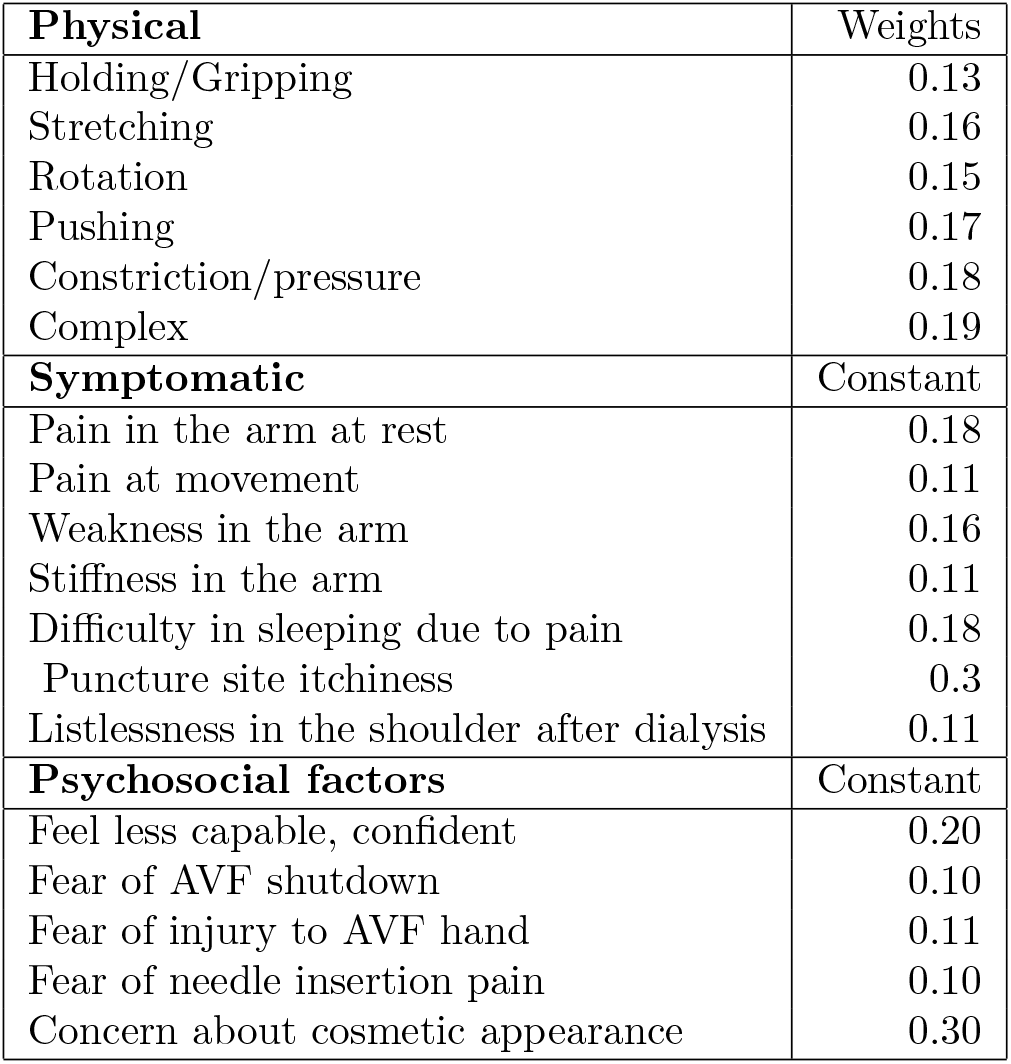
Relative weights of factors within the category

| **Physical** | Weights |
| --- | --- |
| Holding/Gripping | 0.13 |
| Stretching | 0.16 |
| Rotation | 0.15 |
| Pushing | 0.17 |
| Constriction/pressure | 0.18 |
| Complex | 0.19 |
| **Symptomatic** | Constant |
| Pain in the arm at rest | 0.18 |
| Pain at movement | 0.11 |
| Weakness in the arm | 0.16 |
| Stiffness in the arm | 0.11 |
| Difficulty in sleeping due to pain | 0.18 |
| Puncture site itchiness | 0.3 |
| Listlessness in the shoulder after dialysis | 0.11 |
| **Psychosocial factors** | Constant |
| Feel less capable, confident | 0.20 |
| Fear of AVF shutdown | 0.10 |
| Fear of injury to AVF hand | 0.11 |
| Fear of needle insertion pain | 0.10 |
| Concern about cosmetic appearance | 0.30 |

**Table 2:**
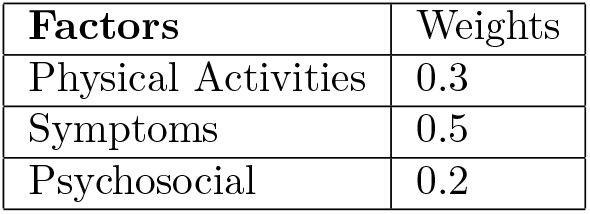
Category weights

| **Factors** | Weights |
| --- | --- |
| Physical Activities | 0.3 |
| Symptoms | 0.5 |
| Psychosocial | 0.2 |

In the relative importance survey, we received 37 responses. The mean importance of each activity was divided by the sum of the mean importance of all activities of a category to compute their weights within that category. The relative importance of each activity (*w*_*i*_) was computed after multiplying the weights of each activity with its category weights.

We have obtained the reduction in “ quality of life” by aggregating the product of relative importance and impact of AVF shown in Table 3 for each activity. Our data analysis showed that dominant arm AVF reduces the QOL (1 *−Q*) of hemodialysis patients by 0.30 (*±* 0.1), whereas the non-dominant arm AVF reduces it by 0.27 (*±* 0.09). We tested the hypothesis (4) using the t-test that the mean reduction of quality of life when AVF constructed on the dominant and non-dominant hand is the same. T-test resulted in *p <* 0.0001, which means that the mean reduction of quality of life for dominant hand fistulas is higher than for the non-dominant hand.

**Table 3:**
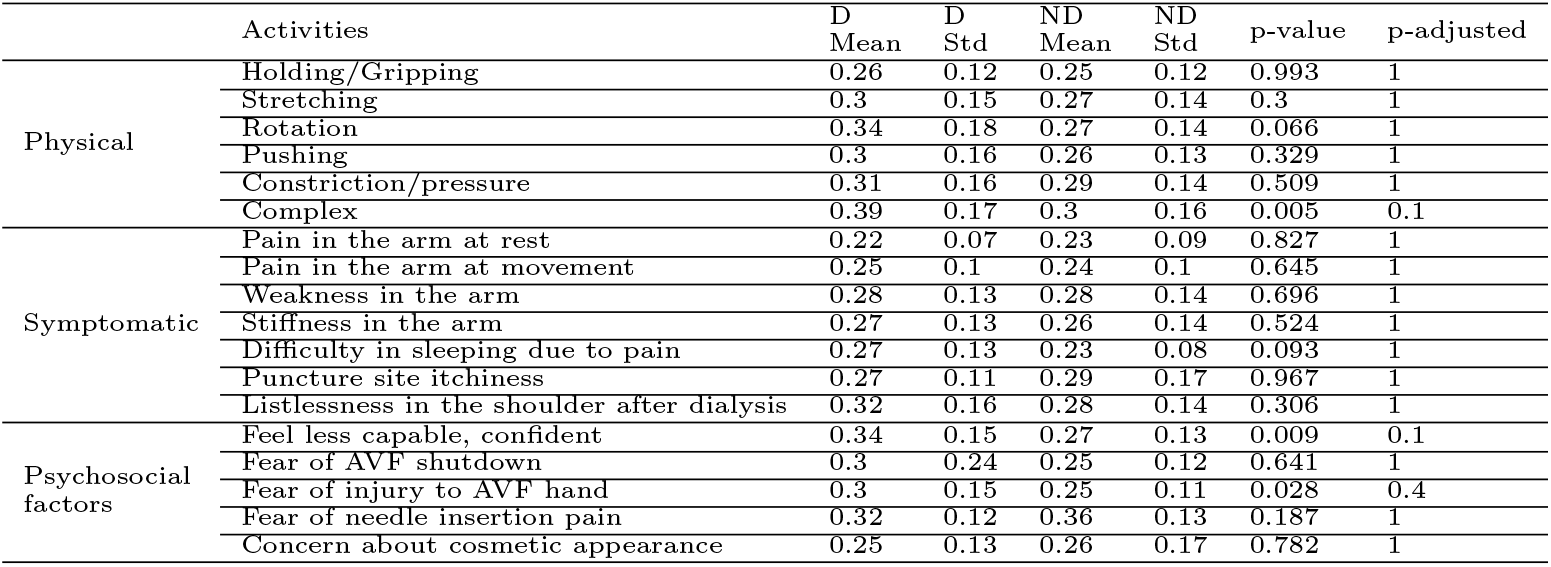
Impact of AVF on each factor.

**Table 4:** Median patency of AVF based on different locations.

| Access Location | Failures (in %) | Median Survival (in months) | Left sided 95% CI |
| --- | --- | --- | --- |
| D Distal | 5/18 (27.8) | 72.4 | 47.6 |
| ND Distal | 23/100 (23) | 91.3 | 73.2 |
| D Proximal | 7/22 (38.9) | 48.43 | 29.07 |
| ND Proximal | 16/85 (18.8) | 84 | 48.2 |

## 6. Observations & Analysis

The dataset was obtained from multiple centers, and four independent distributions were fitted to estimate AVF survival. The event of interest was considered a ‘secondary failure,’ and censoring data included the AVFs that were either still working, patients who were lost to follow-up, and patients who died with functional AVFs. Secondary failure corresponds to long-term failure and is, therefore, appropriate for our decision policy. In our dataset, a total of 51 AVFs had a secondary failure. This event occurred in 12 cases for the dominant arm AVF, and 39 events occurred in the non-dominant arm AVF. The parametric survival functions, listed in section 3.2, were estimated on the above dataset.

The goodness of fit shows that all parametric models have equivalent performance in the data considered in this study. The optimal parameter for exponential for the dominant arm is *λ* = 98.37, whereas for the non-dominant arm is *λ* = 148.63. Fig. (3a) shows the survival plot of the exponential survival function.

The quality-adjusted survival has two terms corresponding to the quality of life and the AVF patency. An AVF patency for an individual patient can be estimated by incorporating the patient’s characteristics. It helped us to propose a patient-specific treatment plan, as in this study, the quality of life is assumed to be independent of the patient. We attempted the multivariate survival analysis of AVFs on the dominant and non-dominant arms by including characteristics such as arm used, patient’s age at the time of fistula construction, diabetes mellitus status, cardiovascular problems, and smoking/tobacco use status. Fig. (3b) shows the log hazard of all these variables, and none of them were statistically significant at a 5% significance level. This means that we do not have statistical evidence of whether the secondary failure of AVF depends on a patient’s characteristics.

The median patency of the dominant arm AVF was 72.4 months (CI^2^: 34.3 - *∞*), and for the non-dominant arm AVF, it was 91.3 months (CI: - 179.67). This was computed using the Kaplan-Meier estimate for the first AVF. All the statistical tests discussed in the section 3.2 show that the dominant arm AVF patency is not statistically different from that of the non-dominant arm AVF patency (*p >* 0.1).

Fig. (4a) shows the quality-adjusted survival of either hand for a period of five years. It is evident from Fig. (4a) that this quality-adjusted survival is always higher for the non-dominant hand.

We showed that the patient’s QOL is affected less when the AVF fistula is constructed on the ND arm and also scores over the dominant arm in terms of AVF patency. This, therefore, indicates that the first AVF should be created on the non-dominant arm to achieve better quality-adjusted AVF survival (*p <* 0.05). Hence, the first and second sequences (discussed in the section 2) can not be optimal for multiple AVF planning. We have tested the hypothesis that the second AVF on the ND arm proximal location has the same hazard ratio as the D distal arm fistula since the hand and the first AVF location are fixed based on the above discussion. We used the Tarone-Ware test to test our hypothesis because both the survival functions were intersecting. We found the p-value between D distal and ND proximal to be 0.58. This indicates that D distal and ND proximal have similar survivals. Figure (4b) shows the AVF survival at different locations. Although patencies of D distal and ND proximal AVFs are not statistically significant, the median patency is higher for ND Proximal as compared to D Distal, as depicted in Table (4). Based on the QOL and patency rates, the third sequence (3) of multiple AVF should be preferred over the fourth sequence (4).

## 7. Conclusion

Given the fact that patients on dialysis currently survive for a very long time, and that their vascular access has a relatively shorter life span, and that there are only a limited number of location options to insert a vascular access in a given patient, this paper addresses the issue of the optimum sequence of locations where such a vascular access can be inserted in these hemodialysis patients. Studies on the quality adjusted vascular access survival favours the distal followed by proximal location on the non-dominant arm, over the dominant arm. Our data showed that vascular access survival statistics were comparable between the two arms and the two locations on each arm. However, quality of life indicators were significantly better when a vascular access was inserted on the non-dominant arm. Our quality assessment is comparable to the detailed study done in [19], but adapted to the patient base in India. It can be revised in other environments as needed.

## Data Availability

All data produced in the present study are available upon reasonable request to the authors.

## Statements and Declarations

### Ethical approval

The institute ethics committee has approved the study.

## Footnotes

1 The Institutional Ethics committee approved this study.

2 CI: Confidence Interval

